## Supplementary material for "Whole-genome mapping of APOBEC mutagenesis in metastatic urothelial carcinoma identifies driver hotspot mutations and a novel mutational signature": Suppementary Figure

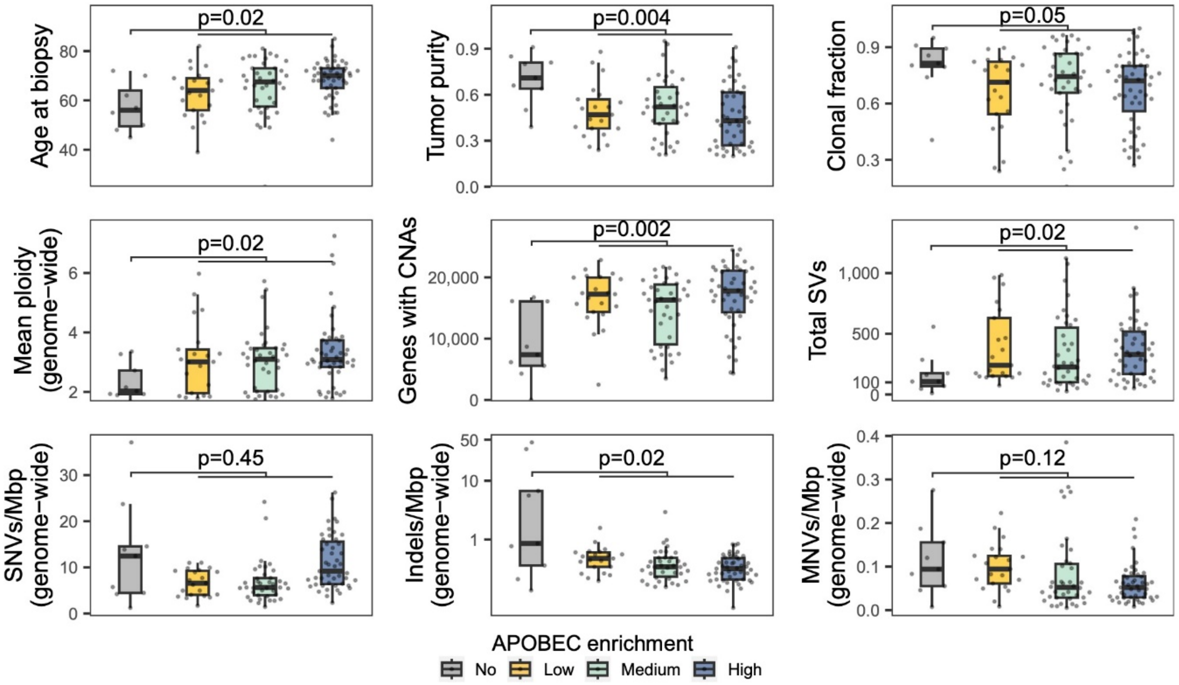


**Figure S1. Distribution of eight genomic features and age according to the level of APOBEC mutagenesis, Related to Figure 1.**

As APOBEC-enriched tumors (low, medium and high) showed similar distributions, they were grouped and compared *vs* non-APOBEC tumors using the Wilcoxon rank-sum test. CNAs = copy number alterations, SVs = structural variants, SNVs = single nucleotide variants, Mbp = megabase-pair, Indels = small insertion/deletions, MNVs = multiple nucleotide variants.

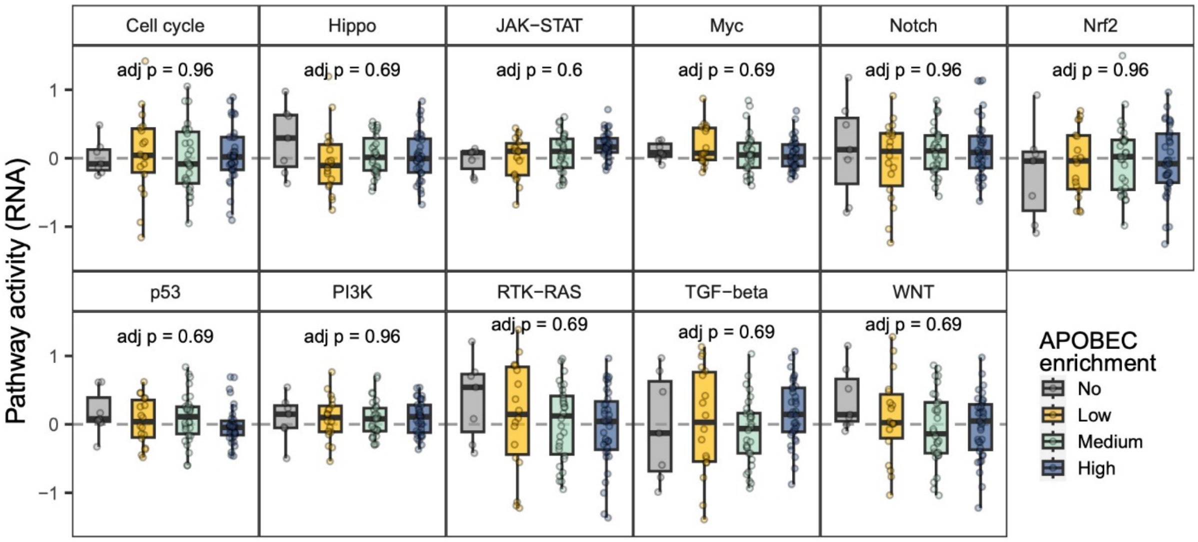


**Figure S2. Pathway activity across APOBEC groups, Related to Figure 1.**
The Kruskal-Wallis test was applied and p-values were adjusted with the Benjamini-Hochberg method. Comparing APOBEC (low + medium + high) vs non-APOBEC tumors using the Wilcoxon rank-sum test showed no difference.

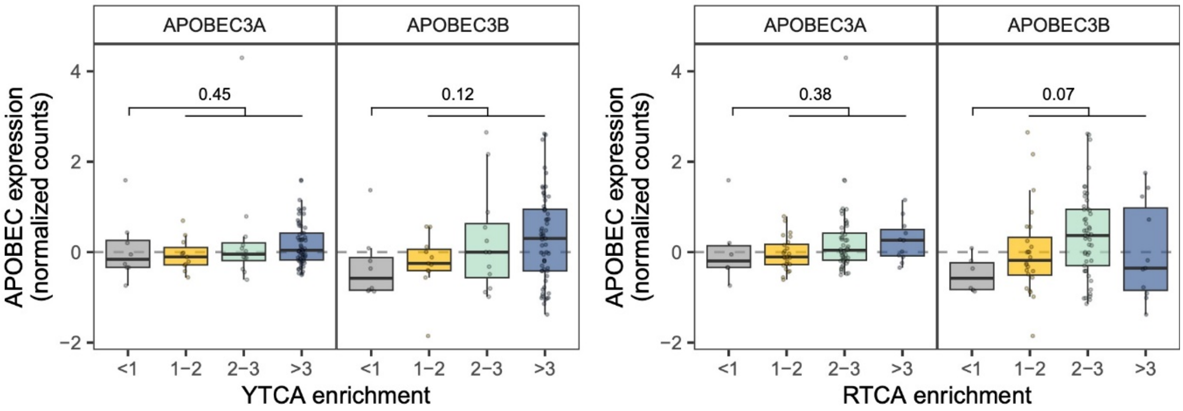


**Figure S3. Expression of *APOBEC3A* and *APOBEC3B* across APOBEC-enriched and non-APOBEC-enriched tumors, Related to Figure 1.**

The Wilcoxon rank-sum test was applied to compare all APOBEC tumors (APOBEC-high, -medium and -low) *vs.* non-APOBEC tumors.

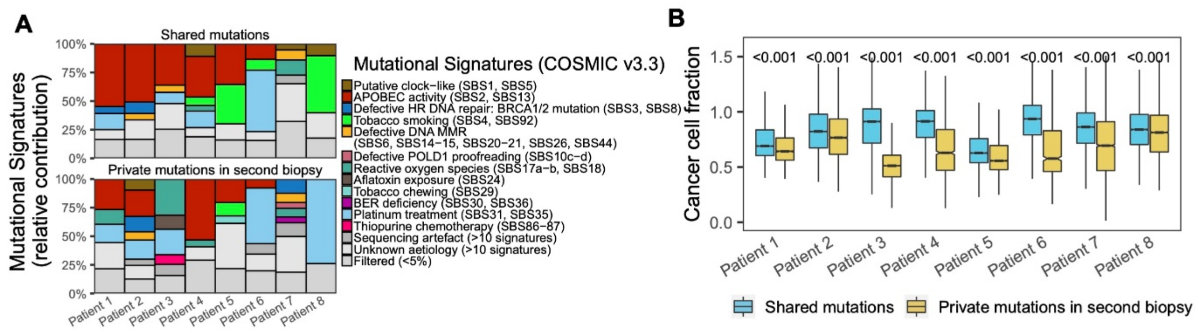


**Figure S4. Tumors with serial biopsies, Related to Figure 1.**

(A) Mutational signatures were calculated from shared single nucleotide variants (SNVs) and from SNVs exclusive to the second biopsy.

(B) Boxplots comparing the cancer cell fraction of somatic mutations from shared SNVs *vs* private SNVs in the second biopsy. In all comparisons, Wilcoxon rank-sum test was applied and p-values were corrected with the Benjamini-Hochberg method.

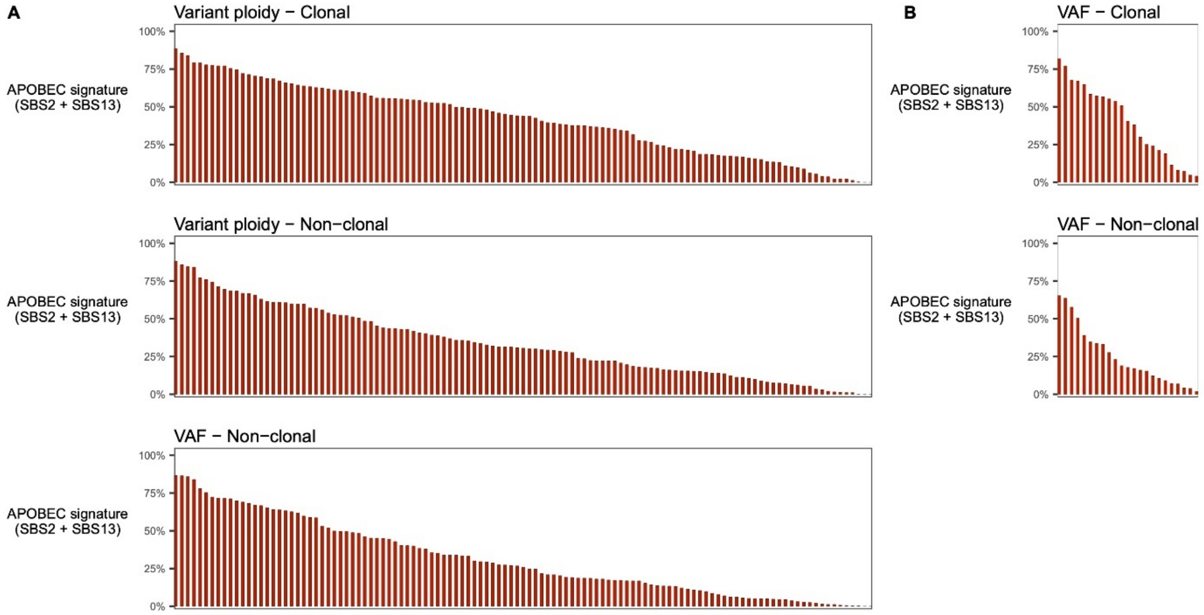


**Figure S5. APOBEC signature within clonal and non-clonal mutations, Related to Figure 1.**

(A) Clonal mutations were estimated using the variant ploidy in metastatic urothelial carcinoma (mUC).

(B) For comparison with 23 primary UC, the variant allele frequency was also used to define non-clonal mutations (VAF<0.125).

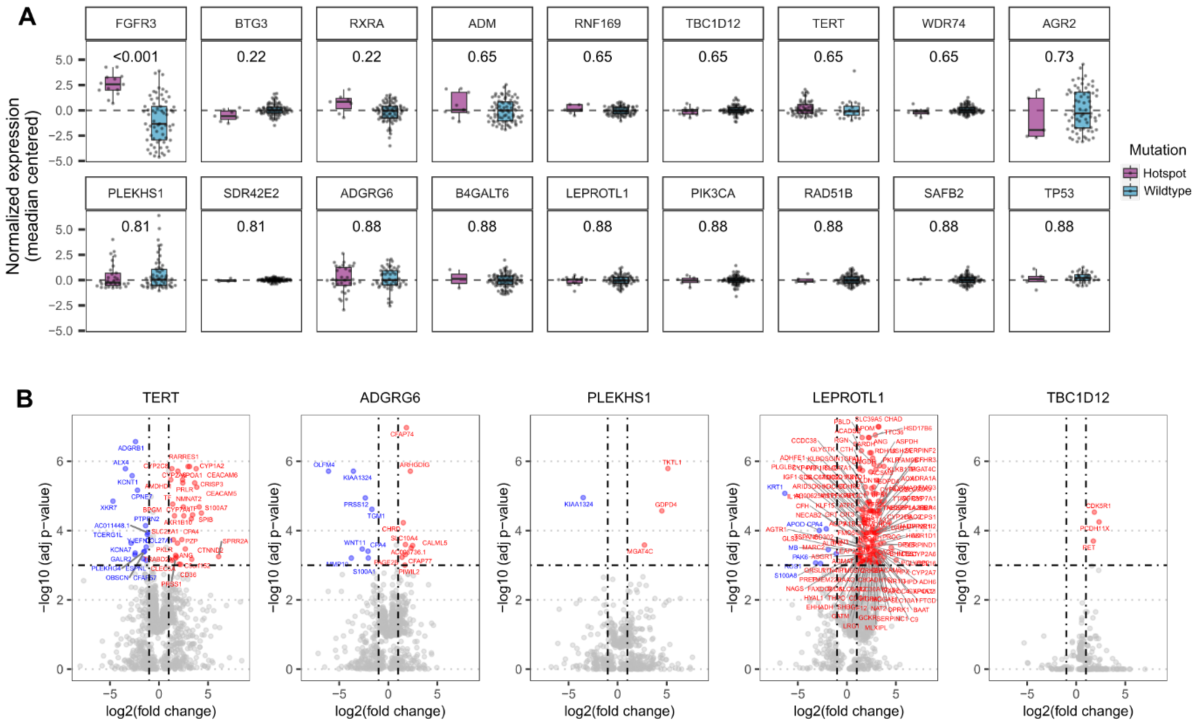


**Figure S6. Transcriptomic effects of hotspot mutations in 90 samples, Related to Figure 3.**

(A) Expression of genes with hotspot mutations *vs* the wild type. Wilcoxon rank-sum test was applied and p-values were Benjamini-Hochberg corrected

(B) Differentially gene expression analysis between tumors with hotspot mutations in specific genes *vs* the wildtype.

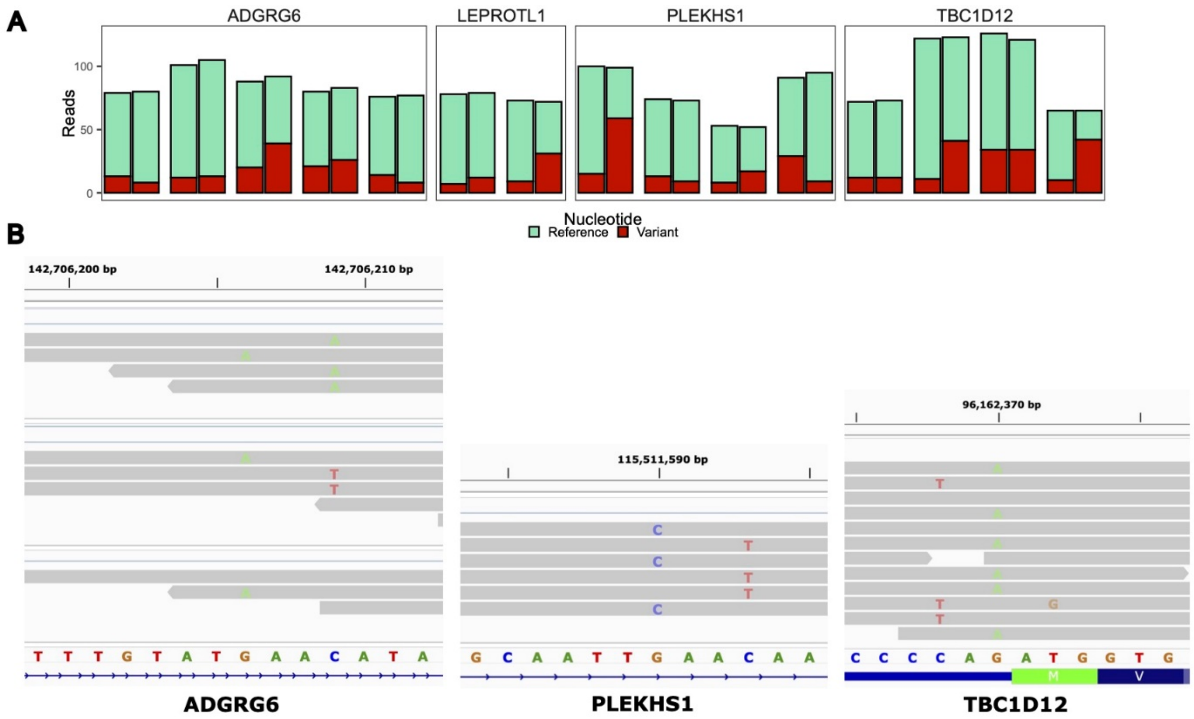


**Figure S7. Patients with co-occurred twin mutations, Related to Figure 4.**

(A) Read counts of the variant and reference alleles of twin mutations grouped per patient.

(B) Raw RNA-sequencing data are shown to interrogate co-occurring twin mutations. In five patients, sufficient coverage was available to show co-occurred twin mutations in *ADGRG6* (n = 3), *PLEKHS1* (n = 1) and *TBC1D12* (n = 1). In all cases, twin mutations did not appear in the same read.

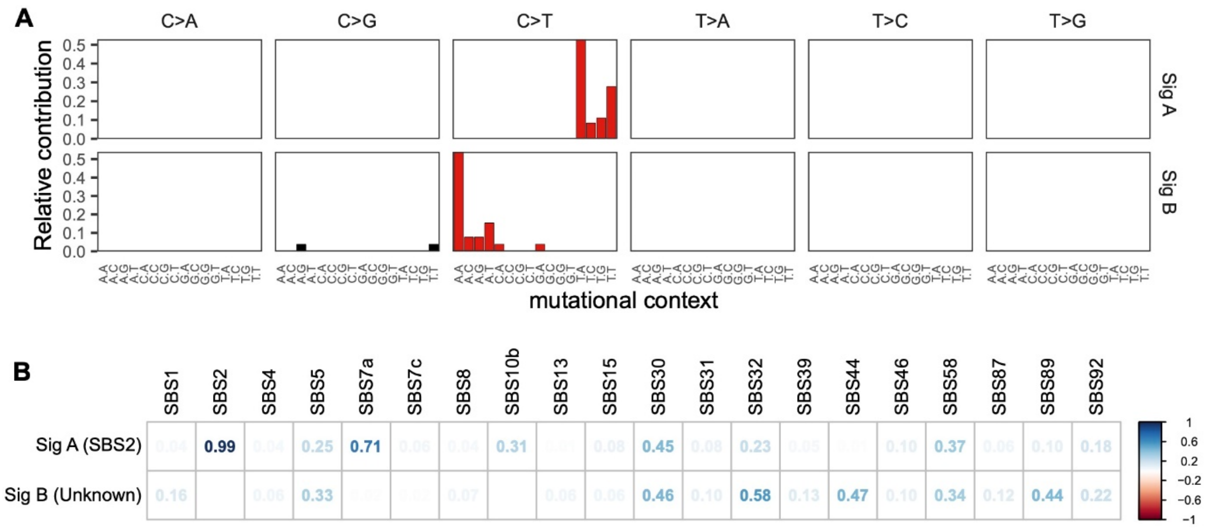


**Figure S8. Mutational signatures of frequent twin mutations that account for 5 or more mutations within a loop, Related to Figure 4.**

(A) Mutational distribution in 96 tri-nucleotide context can be deconvoluted into two signatures (sig A and Sig B).

(B) Cosine similarity of signatures present in twin mutations with the COSMIC signatures.

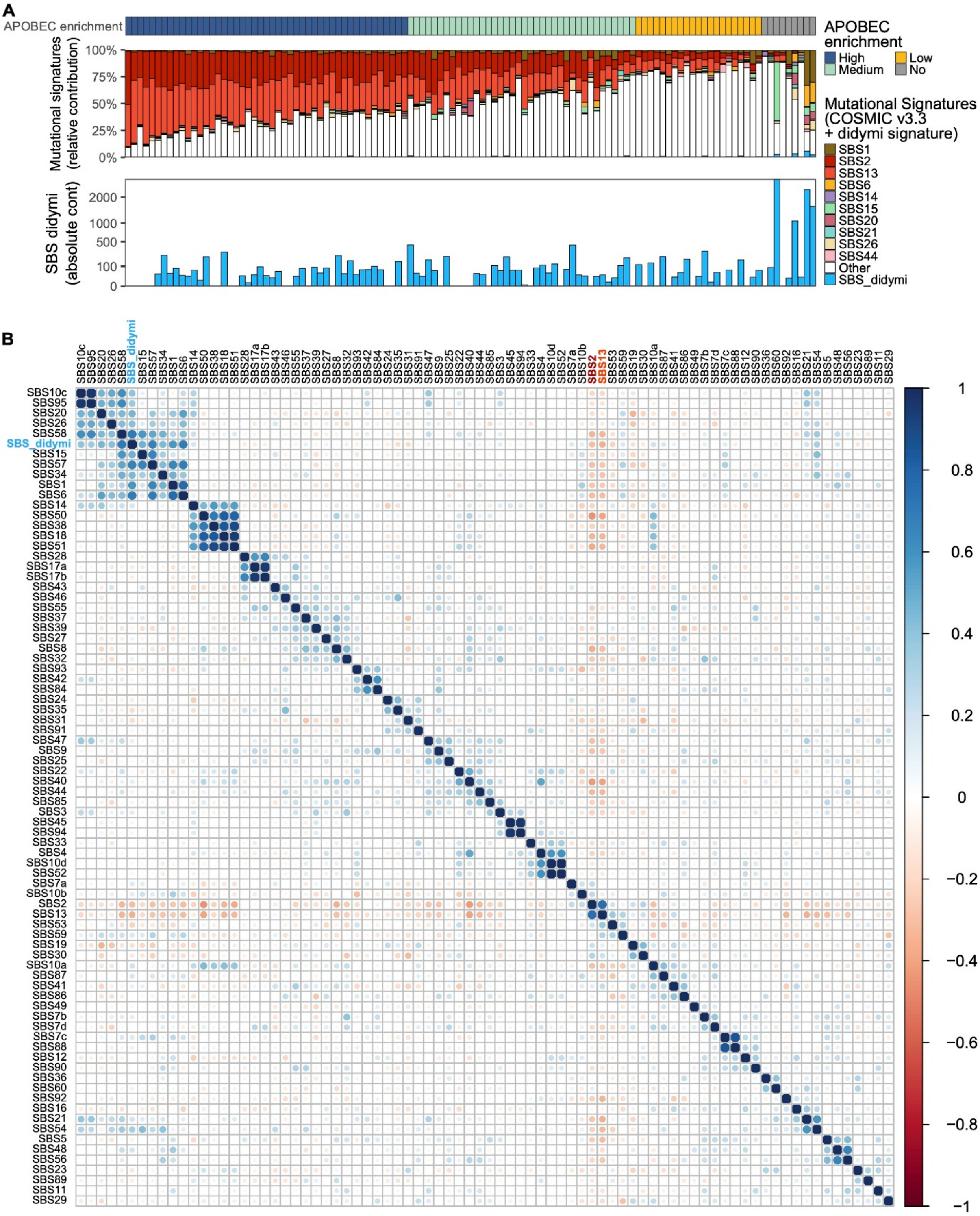


**Figure S9. The prevalence of the ApC signature associated with didymi in 115 metastatic urothelial carcinomas, Related to Figure 4.**

Mutational calling was performed using all COSMIC v3.3 signatures and Sig B from Figure S8 (SBS_didymi).

(A) The relative contribution of mutational signatures stratified by the level of APOBEC mutagenesis and the absolute contribution of the SBS_didymi signature.

(B) Pearson correlation between the relative contribution of all signatures across the whole cohort. The color and size of the circles indicate the correlation value.

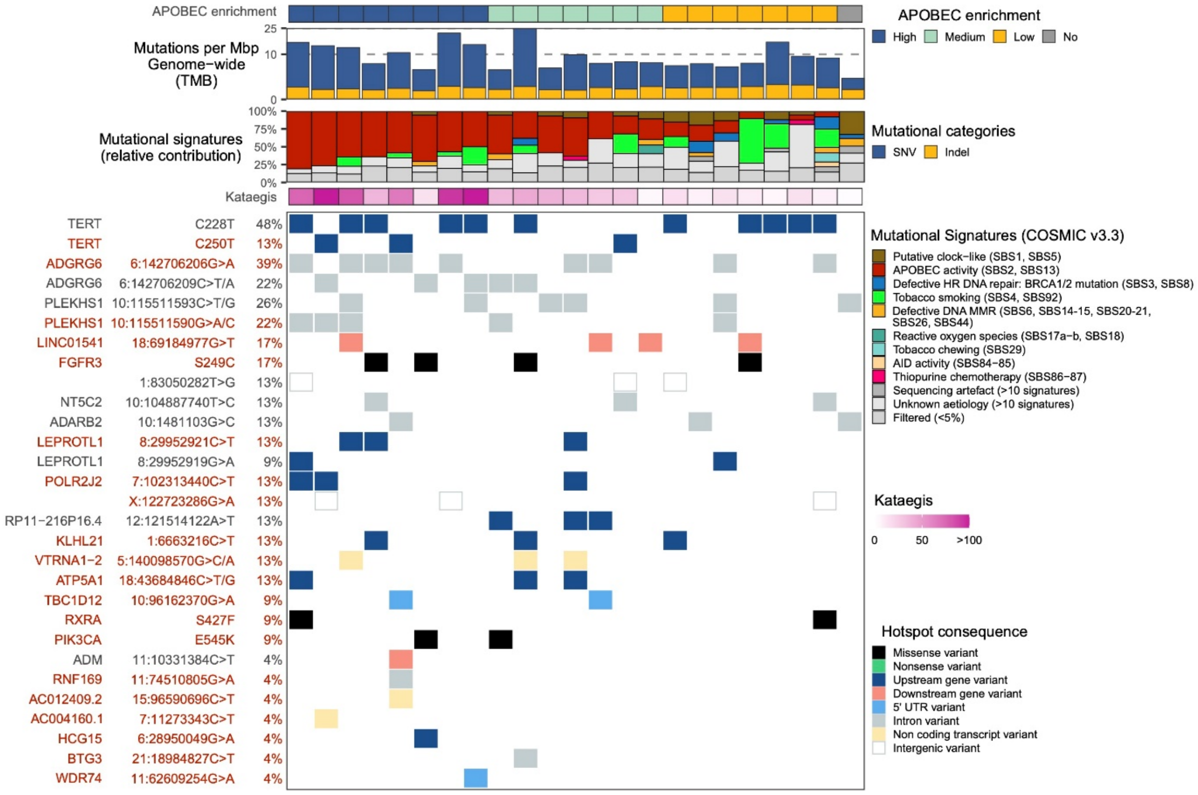


**Figure S10. APOBEC mutagenesis in 23 whole-genomes of primary bladder cancer, Related to Figure 5.**

The genomic features are displayed from top to bottom as follows: APOBEC mutagenesis; genome-wide tumor mutational burden (TMB); mutational signatures grouped by etiology; frequency of kataegis and; the most frequent hotspot mutations, including those found in the metastatic setting.

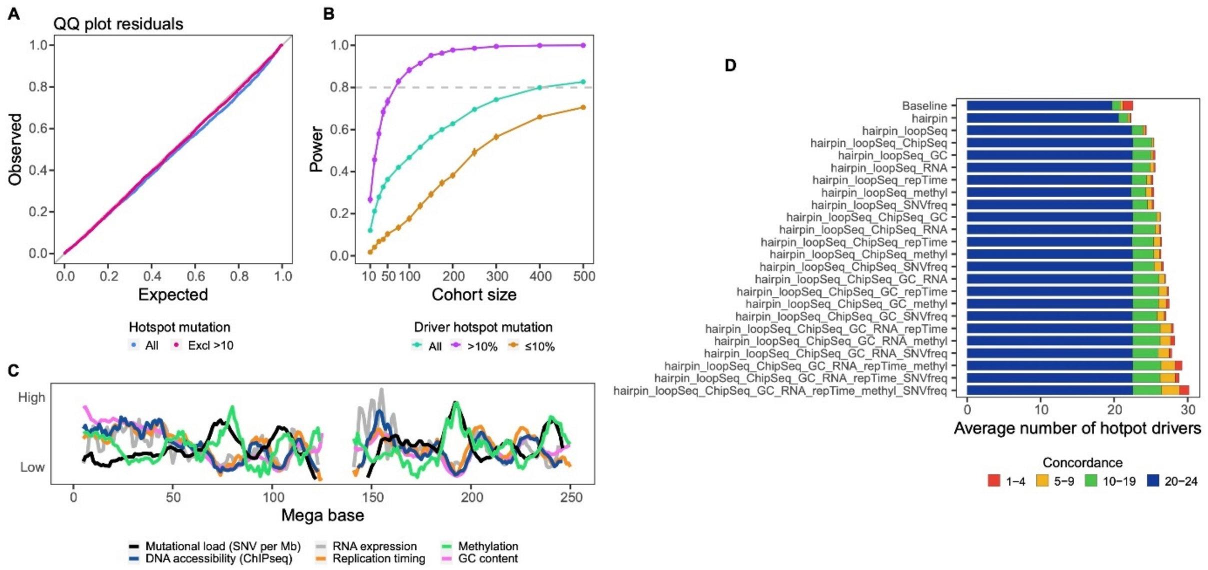


**Figure S11. Performance of the model for detecting driver APOBEC-associated hotspot mutations (ApoHM) in hairpin loops, Related to Figure 5.**

(A) QQ plots of the cumulative probability for all ApoHM (p-value < 0.001, KS-test) and when excluding those present in >10 samples (p-value = 0.19).

(B) Power estimation for the detection of driver ApoHM as a function of cohort size based on simulations of a synthetic genome of metastatic Urothelial Carcinoma.

(C) Variation of genomic features across chromosome 1.

(D) Sensitivity of the model when considering other covariates and their combinations. Concordance indicates the number of models that detected the same ApoHM as a driver.

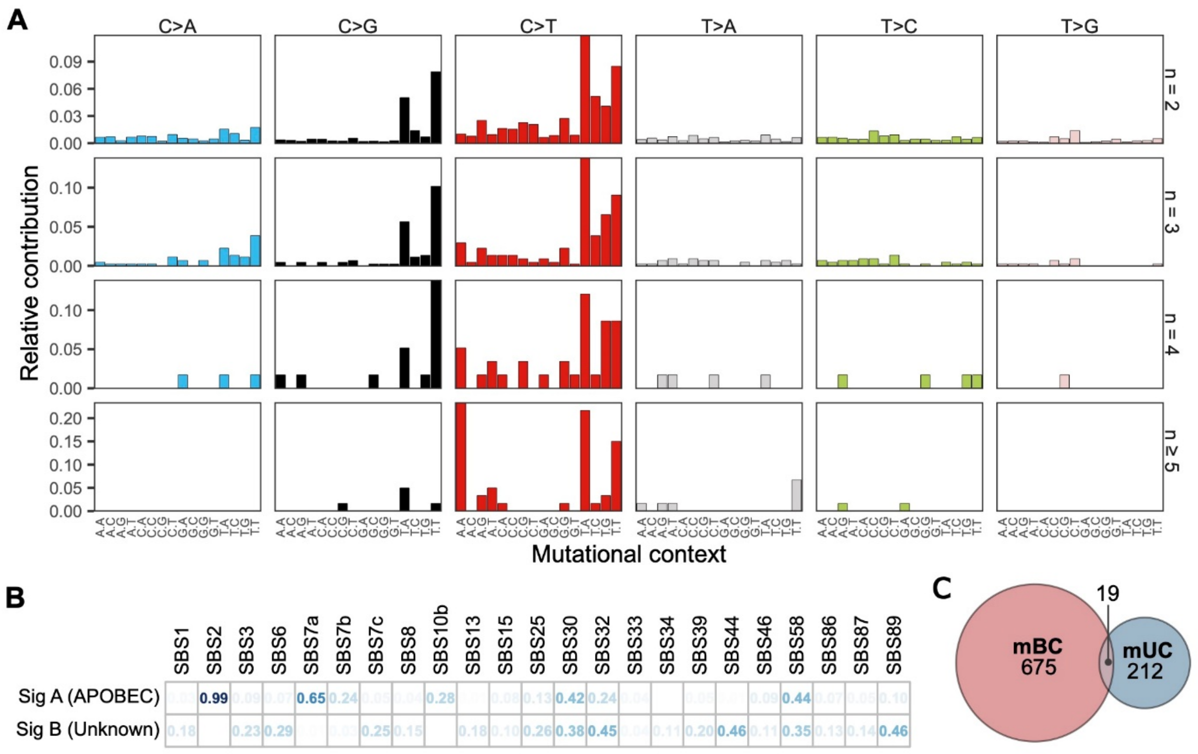


**Figure S12. Twin mutations in metastatic breast cancer, Related to Figure 6.**

(A) Mutational distribution of twin mutations grouped according to the total number of mutations they contributed to the loop (2, 3, 4 or ≥5).

(B) The mutational distribution of frequent twin mutations (n ≥ 5) can be deconvoluted into two signatures and their cosine similarities with the COSMIC signatures are displayed.

(C) Venn diagram of twin mutations classified as didymi identified in metastatic breast cancer (mBC) and metastatic urothelial carcinoma (mUC).
